# Are probiotics and prebiotics safe for use during pregnancy and lactation? A systematic review and meta-analysis

**DOI:** 10.1101/2021.01.19.21250133

**Authors:** Hauna Sheyholislami, Kristin L. Connor

## Abstract

Probiotic and prebiotic products have shown potential health benefits, including for the prevention of adverse pregnancy outcomes. The incidence of adverse effects in pregnant people and their infants associated with probiotic/prebiotic/synbiotic intake, however, remains unclear. The objectives of this study were to evaluate the evidence on adverse effects of maternal probiotic, prebiotic and/or synbiotic supplementation during pregnancy and lactation and interpret the findings to help inform clinical decision-making and care of this population. A systematic review was conducted following PRISMA guidelines. Scientific databases were searched using pre-determined terms, and risk of bias assessments were conducted to determine study quality. Inclusion criteria were English language studies, human studies, access to full-text, and probiotic/prebiotic/synbiotic supplementation to the mother and not the infant. 11/100 eligible studies reported adverse effects and were eligible for inclusion in quantitative analysis, and data were visualised in a GOfER diagram. Probiotic and prebiotic products are safe for use during pregnancy and lactation. One study reported increased risk of vaginal discharge and changes in stool consistency (Relative Risk [95% CI]: 3.67 [1.04, 13.0]) when administering *Lactobacillus rhamnosus* and *L. reuteri*. Adverse effects associated with probiotic and prebiotic use do not pose any serious health concerns to mother or infant. Our findings and knowledge translation visualisations provide healthcare professionals and consumers with information to make evidence-informed decisions about the use of pre- and probiotics.

## Introduction

The use and acceptance of probiotic products is increasing globally due to their documented health benefits[1,2]. Probiotics are live microorganisms that, when administered in adequate amounts, confer a health benefit on the host[3]. Prebiotics, compounds that induce growth or activity of beneficial microorganisms, such as bacteria and fungi[4], and synbiotics, combinations of probiotics and prebiotics[5], are also products that have the ability to influence an individual’s microbiomes[4,5]. In non-pregnant individuals, probiotics have been shown to support a healthy gut and digestive tract, including in the treatment or prevention of *Clostridium difficile*-associated diarrhea[6], irritable bowel syndrome[7], abdominal pain and bloating[8], and necrotizing enterocolitis[9]. Additionally, probiotics may provide health benefits in pregnancy, such as the prevention of gestational diabetes[10], mastitis[11], constipation[12], post-partum depression[13], and growth of Group B *Streptococcus* bacteria[14]. Some probiotic products can alter vaginal and breastmilk microbial composition, and therefore have been used to prevent the recurrence of bacterial vaginosis[15] and support the gut health of newborn infants by influencing the composition of their gut microbiomes[16]. Thus, given their documented and potential therapeutic effects for certain conditions in non-pregnant, pregnant and lactating people, there may be specific situations for which there is a need for probiotics. While probiotic supplementation in pregnant populations has been associated with several health benefits, there is a lack of information on the health benefits of prebiotics and synbiotics in pregnant and lactating people. The potential benefits of these products in non-pregnant populations include reducing plasma cholesterol and insulin concentrations[17], decreasing body weight in overweight individuals[18], improving stool consistency[19], and treating constipation[20].

Despite potential benefits, there is uncertainty about the effectiveness of probiotics, prebiotics and synbiotics for human health, largely because many studies do not consider that the effects are likely strain-, dose- and condition-specific, and an individual’s response to an intervention may be unique, in part determined by their health status, age, and composition of their gut microbiome[21]. Critically, many studies fail to report on adverse effects or do not provide specific details regarding adverse effects, such as timing, duration, and severity of symptoms[22]. Lack of safety data collection and reporting may contribute to unwanted or unexpected health outcomes, which may be of particular concern for vulnerable populations, such as pregnant individuals. Indeed, up-to-date and easy to understand evidence on the safety of probiotic, prebiotic and synbiotic use during pregnancy and lactation is currently limited. This is a critical gap in knowledge, given that many physicians consider probiotics to be safe[23] but want to learn more about them[24,25], and women of childbearing age are highly receptive to taking probiotic-containing products, including to treat gastrointestinal symptoms[26]. Importantly, studies investigating probiotic, prebiotic and synbiotic use on health outcomes often do not present findings in an easy to understand manner for those who can use or be informed by the data, further contributing to ineffective communication of the efficacy and safety of these products.

Whilst previous studies have demonstrated the efficacy of probiotics in pregnancy[27,28], the purpose of this review was to assess their safety. Investigating adverse effect incidence following probiotic, prebiotic or synbiotic supplementation around the time of pregnancy, and synthesizing this evidence in an accessible way, is currently needed to determine whether there are risks that may be associated with their intake for populations that are vulnerable, and to ensure informed decision-making about their potential use to support an individual’s health. This study therefore aimed to 1. systematically synthesize the evidence on adverse effects of maternal probiotic, prebiotic or synbiotic supplementation during pregnancy and during lactation; 2. determine if these interventions are safe and 3. interpret findings to help inform clinical decision-making and care of pre-pregnant, pregnant and lactating people.

## Methods

Our study adhered to the Preferred Reporting Items for Systematic Review and Meta-analysis (PRISMA) guidelines (Supplementary Figure 1 and Supplementary Table 1).

### Information sources and search term

A literature search was conducted between June 5 and June 10, 2020 using the search engines Web of Science, ProQuest, PubMed and CINAHL. Grey literature was not searched, as inclusion of unpublished or non-peer reviewed data may introduce bias^29^. The following search terms were used to retrieve peer-reviewed articles: (pregnancy OR lactation OR prenatal OR pregnant OR preconception) AND (probiotics OR prebiotics OR synbiotics) AND (safety OR effect OR risk OR adverse OR outcome). This search yielded 1793 articles, of which 1519 articles were retained after reference manager duplication (EndNote basic reference manager), and 1185 articles were retained after manual deduplication (Supplementary Figure 1).

### Inclusion criteria

Eligible study designs were randomized control trials (RCTs), case series, controlled clinical trials, case-control studies, cross-sectional studies, and cohort studies. Further inclusion criteria were: 1) English language studies, 2) human studies, 3) access to full-text, and 4) probiotic/prebiotic/synbiotic supplementation to the mother and infant, but not the infant alone.

### Article screening and data collection

A 3-level screening process was performed. Two reviewers defined the strategy and one reviewer executed the search, obtained all articles and performed initial screenings at each level. The second reviewer was consulted at each step of the review process to minimize the likelihood of errors, as per Cochrane guidelines^30^. At level 1, 1185 articles were evaluated based on title and abstract, and articles were screened out if they were not in the English language, human studies, or one of the identified eligible study designs. The remaining 244 articles were carried forward for screening at level 2, where full texts were obtained. Articles were excluded if there was no full-text access and if probiotic/prebiotic/synbiotic supplementation was to the infant and not the mother. Articles were further excluded if the population of interest was not pregnant or lactating people (or if the population did not become pregnant during the study period), if the information provided was insufficient for the requirements of this study, if supplementation was not probiotic, prebiotic or synbiotic-related, and if the article was not a primary research article, leaving 100 studies. Sample size and country of study information were extracted at this level. At level 3, data were extracted from the 100 studies to capture PICO information (patient/population, intervention [timing/duration, strain/supplement type], comparison, and outcome [timing of outcome, statistics related to outcome]. Our primary outcome of interest was adverse effects reporting. Of the 100 eligible studies, 28 studies reported information on adverse events, and of these, 11 studies reported adverse effects. *Adverse events* were identified as unfavorable or harmful outcomes that occur during or after the use of the intervention, but are not necessarily caused by it, whereas *adverse effects* were identified as adverse events for which the causal relationship between the intervention and the event is at least a reasonable possibility^31^. Thus, unfavorable outcomes that are commonly experienced during or after pregnancy, such as pre-eclampsia^32^, were considered adverse events and not adverse effects. Data were obtained for the type, number and timing of adverse effects. Because the studies that reported adverse effects only recruited participants and administered their interventions during or after pregnancy and not before pregnancy, our analyses only include adverse effects that were reported during or after pregnancy. In order to determine the magnitude of the adverse effects reported, the number of each incident in both intervention and non-intervention groups was reported as a risk ratio (RR)^33^. We also noted the primary outcome of interest of the studies under review, and categorized each study based on the primary outcome into 9 groups: pregnancy outcome, maternal metabolic health, maternal microbiome and gastrointestinal health, maternal breast health and breastmilk, maternal mental health, infant metabolic health, infant microbiome and gastrointestinal health, infant allergy and immune health, and infant growth and development (Supplementary Table 2).

### Data synthesis and visualisation

A Graphical Overview for Evidence Reviews (GOfER) diagram^34^ was created to visualise adverse effects reporting and to synthesize key data collected from studies, including timing of study outcome measurements and adverse effect measurements (if applicable), the number of adverse effect-related incidents in intervention and non-intervention groups, risk ratio calculations, intervention type (single factor intervention or multi-factorial intervention), strain type (if applicable), study design (RCT, cohort, case-control or controlled clinical trial), study timeline, primary outcome groups and risk of bias assessment scores. Heatmaps and a world map were created (R Software Version 3.6.1) to represent the quality assessment scores, and the country of study, respectively, for all studies included in level 3. Alluvial diagrams were created (RAWGraphs^35^) to illustrate the relationships between the intervention types used, and the primary outcome groups of the studies. A bubble plot (JMP Software Version 15.1.0) was created to represent the number of studies that reported a specific adverse effect stratified by intervention type. Finally, a funnel plot^36^ was created with the log risk ratio values for each of the reported adverse effects against standard error to represent potential publication bias of all studies included in the meta-analysis.

### Methodological quality assessment

Articles were assessed for methodological quality according to study design using the following scales: the Cochrane Collaboration’s Tool for Assessing Risk of Bias for randomised controlled trials^37^ (n=95), the Newcastle-Ottawa Quality Assessment scale for cohort studies^38^ (n=3) and case-control studies (n=1), and the ROBINS-I tool for clinical trials^39^ (n=1). For RCTs, “other bias” was defined as uptake of the intervention by participants, as non-adherence to the study intervention(s) by the participants may result in performance bias^40^. For cohort and case-control studies, comparability of the exposed/intervention group and not exposed/no intervention group was determined based on whether or not the study’s analyses controlled for 1) other dietary intakes and 2) supplement intake. Dietary intake and other supplement intake were chosen as the comparator variable of the exposed/intervention group to the non-exposed/no intervention group in the quality assessments, as diet exerts one of the largest pressures on the gut microbiome^41^ and thus, may alter one’s response to interventions that target or involve the microbiome. Further, intake of specific foods or supplements may alter the function(s) of prebiotic/probiotic/synbiotic supplementation ^42^. Risk of bias summary scores were calculated for each individual study to determine the overall quality of the study by dividing the number of items with “low risk of bias” by the total number of items included in the tool^43^. Risk of bias summary scores were not calculated for the cohort and case-control studies and clinical trials, as the number of these studies was low (n=5).

### Data analysis

A random effects^44^ meta-analysis was performed on data from studies that reported the incidence of adverse effects in intervention and control groups (n=11). Risk ratio was chosen as an estimate for effect size^44^. Statistical significance was confirmed at α=0.05 and results are presented as risk ratio (95% CI). A summary risk ratio statistic was calculated for all single factor probiotic studies with low risk of bias (n=5). One study (Mirghafourvand et al.) reported adverse effects weekly for four weeks^12^. The final week of the study was chosen as the clinically important timepoint and was used to calculate the risk ratio of the study, as per Cochrane guidelines^45^. This is because most of the symptoms reported following probiotic use begin early and taper off upon continuous ingestion of these products^46^. Additionally, this method allowed for consistent analysis of adverse effects across studies, where in all remaining studies, adverse effects were captured at the end of the intervention period. In another study (Rautava et al.) there were two intervention groups and a single control group. We dealt with this by evenly dividing the number of participants and adverse effects in the control group in half to serve as two separate comparison groups for each intervention group, as per Cochrane guidelines^47^. Finally, absolute risk difference (ARD) was chosen as a secondary effect estimate^48^ for studies included in the meta-analysis (n=11) (Supplementary Table 3). Positive ARD suggests decreased risk of an adverse effect, whereas negative ARD suggests increased risk of an adverse effect.

## Results

### Study location, demographics and design: 70 studies under initial review

The articles under review included data from 25 countries (Supplementary Figure 2). Fifty-four percent of studies were from Europe (n=54), 28% from Asia (n=28), 13% from Oceania (n=13), and the remaining 5% from North America, South America and Africa (n=5). Finland had the highest representation in studies under review (n=29).

For the 100 studies under initial review, 95 were RCTs, 3 were cohort studies, 1 was a case-control study and 1 was a controlled clinical trial. Other study types were not represented at the data extraction level as they were excluded based on predefined eligibility criteria. Of the 100 studies, 49% (n=49), including all three of the observational cohort studies, did not include any information related to safety or adverse events/effects, 23% (n=23) deemed their intervention to be safe (stated that no adverse events/effects were reported or stated that the intervention was safe, without showing safety data), 28% (n=28) reported adverse events, and of these 28 studies, 39% (n=11) reported adverse effects. Eighty percent (n=80) of the 100 studies used a single factor intervention (probiotics [n=68], prebiotics [n=2], synbiotics [n=7], questionnaire [n=2], or fermented foods containing prebiotic [n=1]), and 20% (n=20) used a multi-factorial intervention (diet and probiotic [n=13], multiple probiotic [n=2], vitamin D and probiotic [n=1], complex nutritional supplement and probiotic [n=1], antenatal care and probiotic [n=1], fish oil and probiotic [n=1]). All 11 studies included in the GOfER diagram that reported on adverse effects were randomized controlled trials. Of the 11 studies, 6 included single factor interventions (probiotics [n=5] and prebiotics [n=1]), and 5 included multi-factorial interventions (diet+probiotic [n=1], multiple probiotics [n=2], nutritional supplement+probiotic [n=1], and fish oil+probiotic [n=1]) (Figure 3A).

### Methodological quality assessments

Among the RCTs under review (n=95), risk of bias assessment summary scores for each individual study showed that 62.1% (n=59) of studies had overall low risk of bias, 25.3% (n=24) of studies had overall unclear/moderate risk of bias, and 12.6% (n=12) of studies had overall high risk of bias (Figure 1). The cohort studies (n=3) largely met all criteria of the quality assessment tool, with the exception of controlling for other dietary intakes category (n=1), controlling for supplement intake category (n=1), and adequacy of follow up category (n=1) (Figure 1). The case-control study met most criteria of the quality assessment tool, with the exception of selection of controls and controlling for pre-identified confounders. The controlled clinical trial also met most criteria, but did not control for dietary or supplement intake, and many participants were lost to follow up in the control group.

**Fig. 1.**
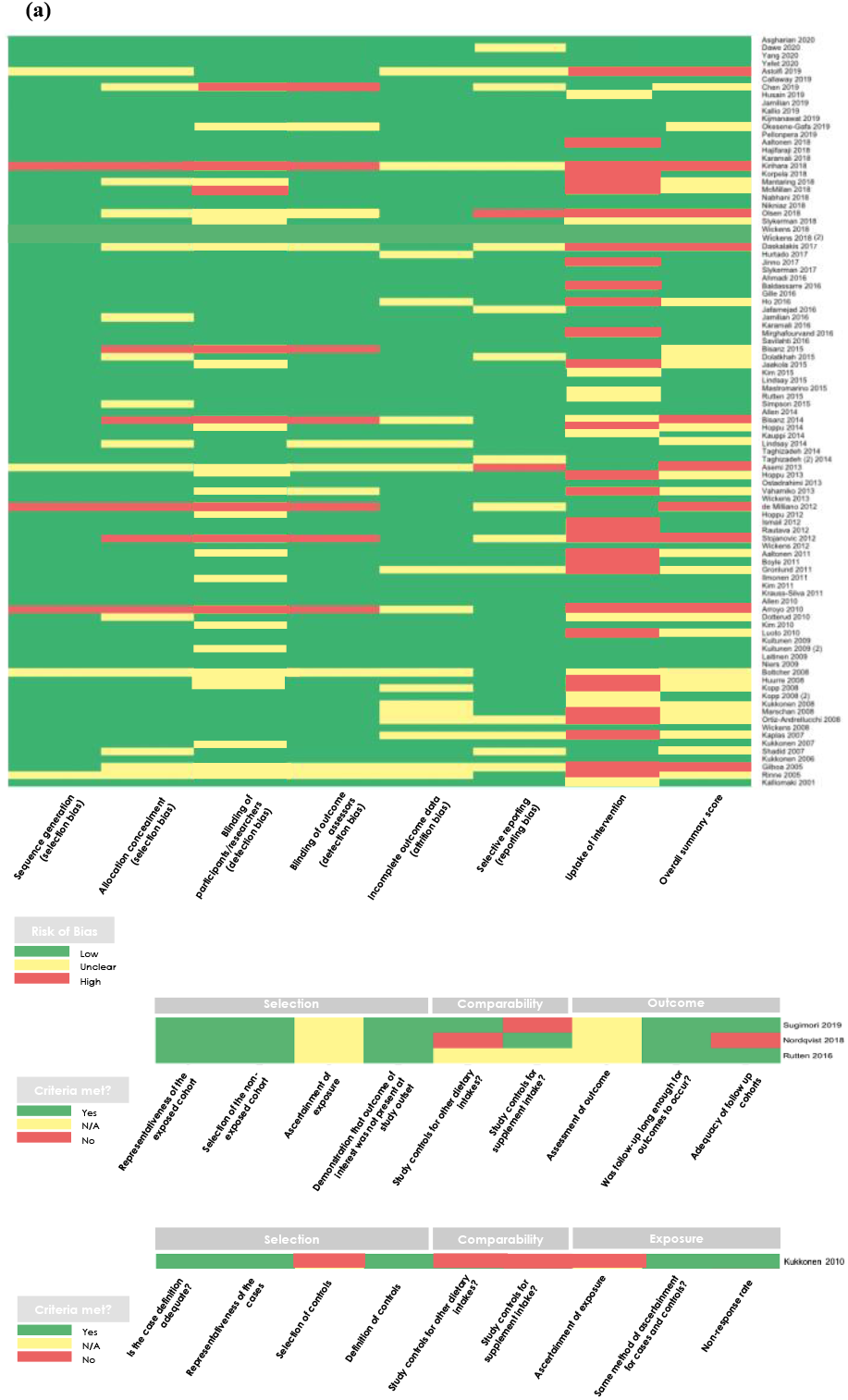
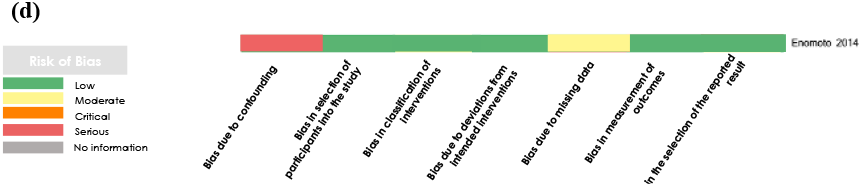
Heatmap of methodological quality assessment scores for (a) randomized controlled trials (n=95), (b) cohort studies (n=3), (c) case-control studies (n=1) and (d) controlled clinical trials (n=1). For RCTs, green = low risk of bias, yellow = unclear risk of bias, red = high risk of bias. For cohort and case-control studies, green = criteria met, yellow = N/A, and red = criteria not met. For controlled clinical trials, green = low risk of bias, yellow = moderate risk of bias, orange = critical risk of bias, red = serious risk of bias, and grey = no information.

### Characteristics of studies that reported on adverse effects

In total, 20 different adverse effects were reported in 11 studies (Figure 2). The most frequently reported adverse effects were gastrointestinal symptoms (type of symptom undefined; n=3), nausea (n=3), diarrhea (n=4) and constipation or bloating (n=3). The majority of the adverse effects were documented in mothers (n=18, vs. n=2 documented in the infant). Of the 20 adverse effect types reported, 16 were associated with probiotic intake, one was associated with prebiotic intake and nine were associated with multi-factorial interventions that included probiotic supplementation (Figure 2). The duration of the interventions varied from the first trimester of pregnancy to one year postpartum depending on the study, with the exception of one study which did not provide any information on the start or end of the intervention^49^. Of the 11 studies included in the GOfER analysis (Supplementary Table 4), only two studies reported the timing of the adverse effect, whereas the other nine studies did not report any timing information (Figure 4). Further, while seven studies were conducted in healthy pregnant individuals, the other four studies were conducted in pregnant individuals with underlying health conditions. These health conditions included lactational mastitis, constipation, overweight/obesity, and intermediate-degree infections/asymptomatic bacterial vaginosis^12,50-52^.

**Fig 2.**
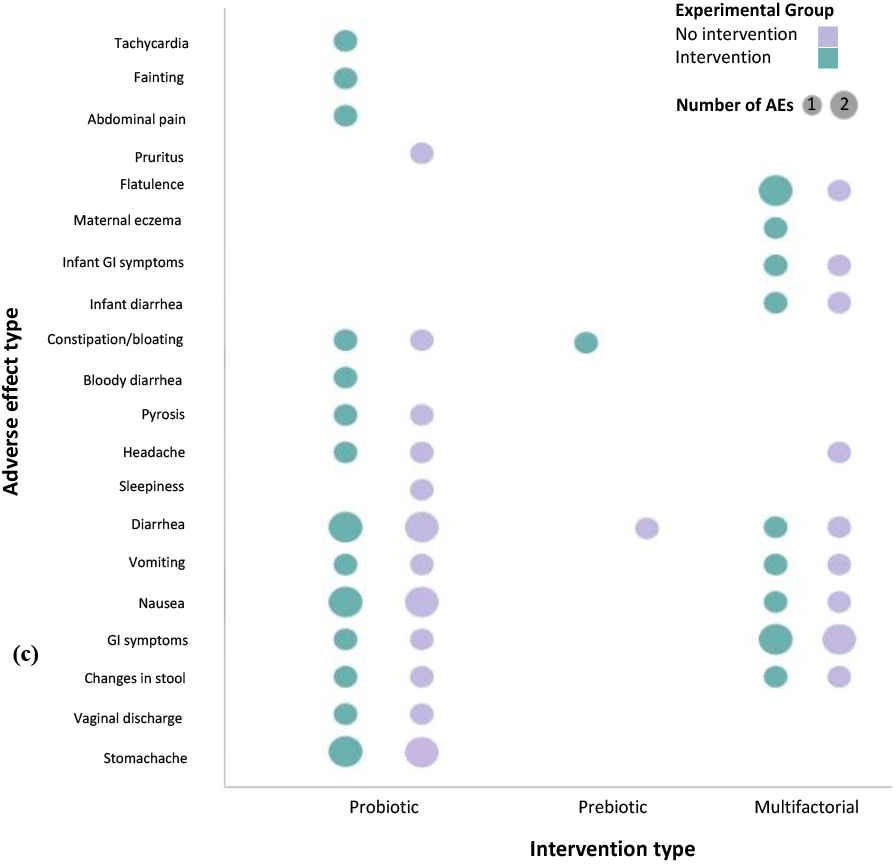
Bubble plot of type and number of adverse effects stratified by intervention type. Size of bubble represents the number of times the given adverse effect was reported for the corresponding intervention across all studies.

**Fig. 3.**
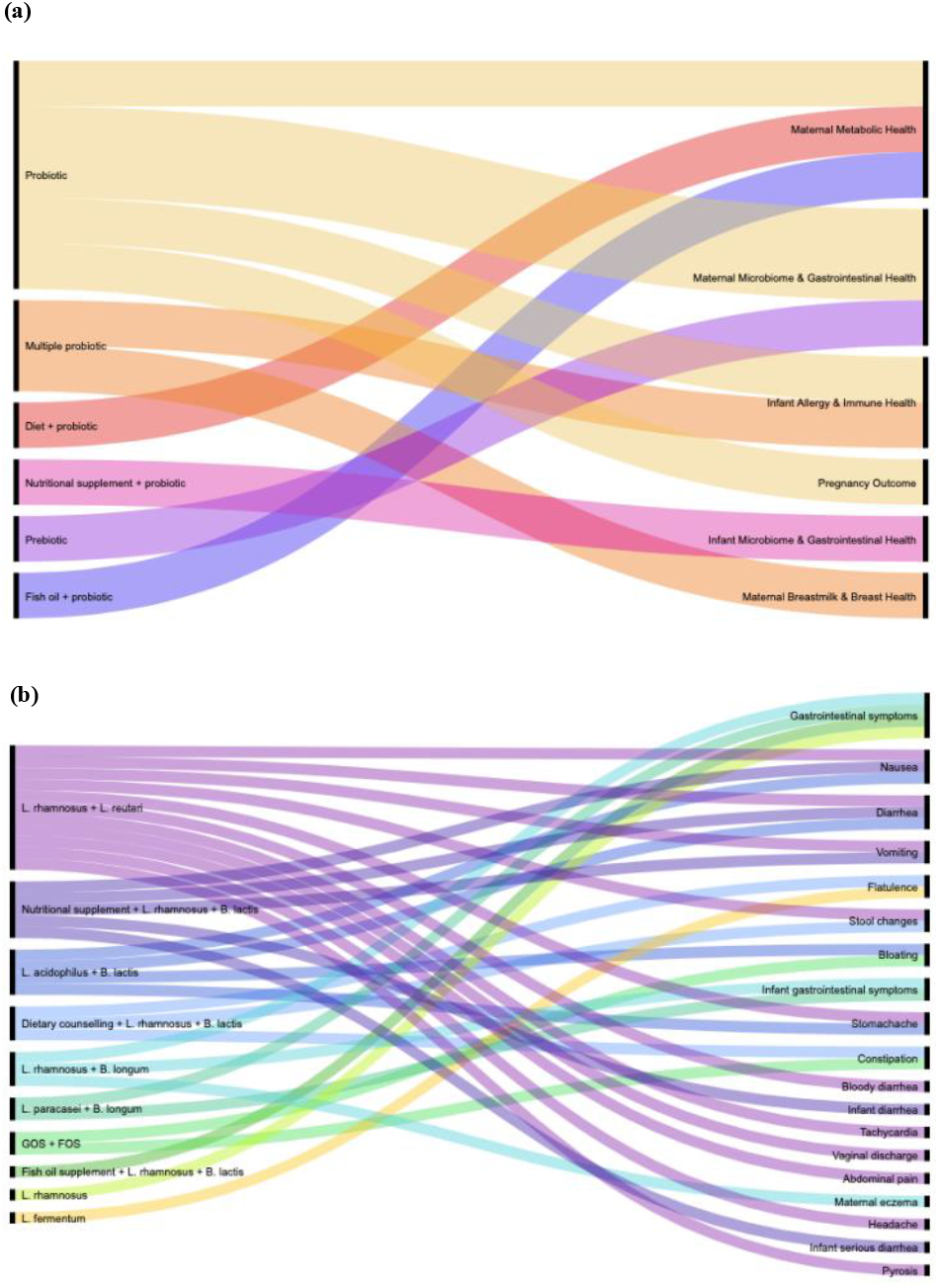
Alluvial diagrams of A. general intervention type and primary outcome group and B. specific intervention (including probiotic products, prebiotic product, and multi-factorial interventions) and adverse effect type (11 studies with reported adverse effects captured).

**Fig. 4.**
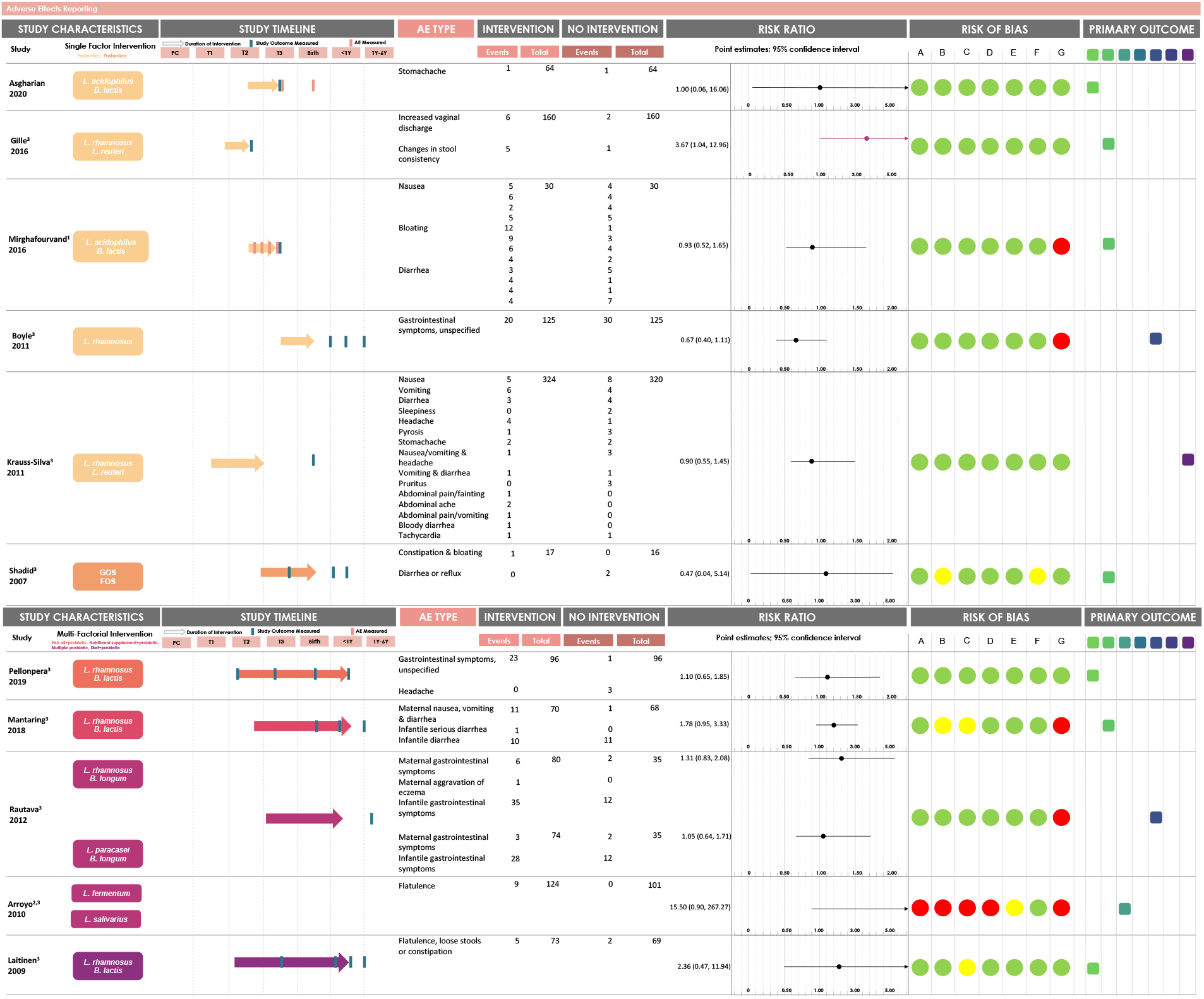
Summary of studies with reported adverse effects. Study characteristics, intervention type, study timeline, number of adverse effect events in each group and major findings are summarized. Risk ratios (RR) are calculated using the number of events in each experimental group. Red point estimates indicate significant RR. Risk of bias was determined for: A. sequence generation, B. allocation concealment, C. blinding of participants/researchers, D. blinding of outcome assessors, E. selective outcome reporting, F. incomplete outcome data, and G. uptake of intervention. 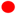 = high risk of bias, 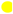 = unclear risk of bias, 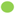 =low risk of bias. Primary outcome groups for each study are shown: 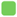 = maternal and infant metabolic health; 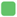 = maternal and infant microbiome & gastrointestinal health; 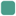 = maternal breastmilk and breast health; 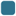 = maternal mental health; 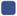 = infant allergy & immune health; 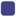 = infant growth & behaviour; 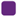 = pregnancy outcome. PC = Preconception. T1, T2, T3 = First, second, or third trimester of pregnancy. <1Y = From birth to 1 year of infant’s age. 1Y-6Y = From 1 year to 6 years of infant’s age. AE = Adverse effect. GOS = Galactooligosaccharide. FOS = Fructooligosaccharide. ^1^Events shown under each adverse effect represent the number of events reported at each week of intervention, over the 4 week intervention period. Risk ratio calculated based on week 4 values. ^2^Does not specify the start time of the intervention. Study duration was 21 days. ^3^Most studies did not indicate when the adverse effect was reported

Various intervention products were used in the studies that reported adverse effects. The probiotic species administered were *Lactobacillus rhamnosus* (n=7), *L. reuteri* (n=2), *L. acidophilus* (n=2), *Bifidobacterium lactis* (n=5), *B. longum* (n=1), *L. paracasei* (n=1), *L. fermentum* (n=1), and *L. salivarius* (n=1). The prebiotic carbohydrates administered were galacto-oligosaccharides (GOS) and fructo-oligosaccharides (FOS) (Figure 3B). Synbiotic products were not administered as an intervention in any of the studies that reported on adverse effects.

### Probiotics are safe for use during pregnancy with minimal reported adverse effects

Of the six studies that used either probiotics or prebiotics alone as an intervention, one study (Gille et al.) reported a statistically significant increased risk of an adverse effect in the intervention group: maternal intake of a probiotic containing *L. rhamnosus* and *L. reuteri* for eight weeks during the first and second trimesters of pregnancy was associated with an increased risk of increased vaginal discharge and changes in stool consistency (RR: 3.67, [1.04, 12.96], Figure 4; ARD: -5.00 [-9.44, -0.55], Supplementary Table 3)^53^. The methodological quality assessment summary score suggests that this study demonstrated an overall low risk of bias (Figure 1). In the study that reported adverse effects over four weeks (Mirghafourvand et al.), there was no statistically significant increased risk of an adverse effect in the intervention group in the fourth week of the intervention (our chosen clinically important timepoint for analysis). However, the number of adverse effect-related events in the intervention group of this study is much higher during the first week of the intervention, which is expected, given the likely mechanism of action of probiotics within the body^46^. Analysis of methodological quality assessment suggests that this study also showed an overall low risk of bias (Figure 1). The summary risk ratio statistic of all probiotic-based single factor intervention studies with low risk of bias (n=5) did not suggest an increased risk of adverse effect incidence associated with supplementation (RR: 0.93 [0.51, 1.71]). Of the five articles that used multi-factorial interventions, none of the studies showed a significantly increased risk ofany adverse effect associated with multi-factorial probiotic-related supplementation (Figure 4).

Finally, low to moderate heterogeneity of the observed effect sizes across the studies was found (I^2^=37.38%). To examine this heterogeneity, an Egger regression analysis was conducted. Visual inspection of the funnel plot showed little asymmetry for studies included in the meta-analysis, and no publication bias was found across the included studies (Begg’s test, p=0.217; Supplementary Figure 3), suggesting that the studies included in our meta-analysis are a representative sample of the available evidence.

## Discussion

Here we describe the first systematic review and meta-analysis to report on adverse effects in individuals taking probiotic, prebiotic or synbiotic supplements during or after pregnancy and during lactation. We present these findings in an easy-to-understand manner to equip knowledge users to make evidence-informed decisions about the potential risks and documented benefits associated with probiotic and prebiotic intake for pre-pregnant, pregnant and lactating people. We found no mortality or serious adverse effects associated with intake of probiotics, prebiotics or multi-factorial interventions including probiotics in pregnant people. Twenty different adverse effects were reported in 11 studies that used probiotics, prebiotics or probiotic-related multi-factorial interventions. Most adverse effects were related to gastrointestinal health in mothers during the third trimester of pregnancy. Maternal intake of a probiotic product alone was associated with increased risk of increased vaginal discharge and changes in stool consistency in one study, although the increased risk of these adverse effects was minimal. Our data suggest that supplementation with probiotic and prebiotic products is relatively safe for use during and after pregnancy and during lactation and is not associated with any serious health outcomes in the mother or infant.

Our evidence synthesis revealed that most reported adverse effects were related to gastrointestinal health, and the majority were reported in the mother. In non-pregnant populations, adverse effects related to probiotic use have been reported, and include gastrointestinal side effects^22^ such as abdominal cramping, nausea, soft stools, flatulence and taste disturbance. These gastrointestinal symptoms are consistent with those documented in our study and suggest that presentation of these adverse effects following intake of some probiotics is not specific to pregnant individuals. Further, while experiencing gastrointestinal-related adverse effects such as nausea, diarrhea, or vomiting during pregnancy may cause discomfort, gastrointestinal symptoms are common and expected during pregnancy^54^, and our analysis suggests that these effects are not increased due to consumption of probiotics or prebiotics. The most frequent gastrointestinal conditions in pregnancy are nausea and vomiting, which affect 50-80% of women^54^, followed by gastroesophageal reflux disease and constipation^54^. Although our meta-analysis showed slight increased risk of increased vaginal discharge and changes in stool consistency, these conditions can usually be managed with lifestyle and dietary modifications, and without the use of medications, under guidance of a physician. There are probiotic products with documented clinical benefits in pregnancy, such as preventing or treating gestational diabetes^10^, mastitis^11^, preterm birth^55^ and infantile atopic dermatitis^56^. Probiotic supplementation in pregnancy has also been associated with improved glucose metabolism^27^, reduced inflammation^57^ and decreased risk of infection^58^. Therefore, these products may contribute to an overall improved health status for pre-pregnant, pregnant and postpartum patients and their children in specific situations, and their benefits may outweigh the documented minimal risks. Nevertheless, it must be acknowledged that some women may not wish to experience the adverse effects associated with these products for long periods of time, especially if the symptom onset is severe.

Reproductive aged women are one of the most common groups to take probiotics^26^, and the most common consumer concerns regarding their use are related to potential side effects and efficacy^1,2^. Additionally, many individuals are unaware of prebiotic products and how they could potentially function to benefit health^2^. Individuals may therefore turn to their healthcare providers for information on the plausible health benefits and safety of these products, especially in countries where product information on the packaging is prohibited^59^. Yet, studies show that many healthcare professionals only have a medium understanding of what probiotics are and how they work, although they believe probiotics are somewhat beneficial to health and are not harmful^24,60^. This suggests that there is a need for knowledge dissemination tools to summarise and translate efficacy and safety information of probiotics and prebiotics to consumers and healthcare providers. Indeed, most healthcare professionals want to learn more about these products^24,25^. Our data visualisations, including the GOfER diagram, provide this information in an accessible manner, empowering the individual and care provider to make evidence-informed decisions for maternal-child health. Our up-to-date analysis of the safety of probiotic and prebiotic use during pregnancy and lactation and assessment of study quality is especially valuable given the prior lack of safety data and the number of studies with inadequate design and/or reporting. Our study complements critical existing efforts to translate scientific evidence on probiotic products into clinically relevant information that can be used to make evidence-informed clinical decisions for both healthcare providers and their patients^61^.

The mechanism underlying adverse effect occurrence following probiotic and prebiotic administration is not completely clear. Most of the studies we reviewed that reported adverse effects used combinations of probiotic strains or prebiotics, making it difficult to ascertain whether the adverse effects occurred because of one strain alone, or complex interventions. Additionally, the timing and duration of intervention administration varied between each study, ranging from the first trimester of pregnancy, to one year postpartum. To better understand the factors contributing to adverse effects associated with probiotic intake, future research should assess individual strain, dose- and timing-specific effects on safety outcomes in pregnancy and postpartum. Additionally, none of the studies reporting on safety outcomes administered synbiotics, suggesting that there is a need for research on, and reporting of, adverse effects related to synbiotic intake, and related to multi-factorial interventions during pregnancy and lactation.

Despite that our evidence review fills a major knowledge gap on safety, there are limitations to some of the studies that were captured. Firstly, while more than half of the RCT studies included in our analysis had an overall low risk of bias, 37.9% had moderate or high risk of bias. Secondly, of the 11 studies that reported adverse effects, the majority did not note the timing or duration of these adverse effects, along with the severity of the specified symptoms, which is concerning as this information is critical to understanding the overall nature of these adverse effects and how they may affect individuals who consume them. Thirdly, less than half of these 11 studies were conducted in pregnant populations with underlying health conditions, including lactational mastitis, bacterial vaginosis, overweight or obesity and history of allergic disease. Current research in non-pregnant populations show that an individual’s gut microbial composition may be influenced by the presence of underlying health conditions, such as inflammatory bowel disease, irritable bowel syndrome, obesity, type 2 diabetes and atopy^62^. Further, an individual’s gut microbiome can determine how drugs are metabolized, which varies on a person-to-person basis^63,64^. Because these underlying health conditions can alter an individual’s gut microbiome, and that probiotic-gut microbiome interactions, like drug-gut microbiome interactions, are likely highly individualized, it is plausible that the efficacy and adverse effects of a pro- or prebiotic intervention will in part be determined by host health status and gut microbiome composition and function. For this reason, a better understanding of the safety of probiotics and prebiotics is not only needed for healthy pregnant and lactating individuals, but also those with underlying health conditions. Notably, animal models may provide insight into the mechanisms through which probiotics, prebiotics and synbiotics may act to elicit outcomes and adverse effects, although these studies are usually focused on the efficacy of interventions, not safety, and rarely report morbidity and mortality data^65,66^. Thus, future animal studies should also report safety outcomes and thoroughly explore adverse effects associated with probiotic, prebiotic and synbiotic supplementation. Lastly, there are limitations to our review. Despite that we searched multiple databases, the possibility exists that we may not have captured all studies that used probiotic/prebiotic/synbiotic interventions in our target populations and reported adverse effects due to lack of full-text access or absence of grey literature. This may include studies that exist in the Cochrane database that were not also indexed in our pre-defined search databases, although our meta-analysis suggests that included studies are a representative sample of the available evidence.

## 5. Conclusions

The global probiotics market size was estimated to be USD 51.12 billion in 2019^67^, and will likely demonstrate continued growth. While this increase is mostly driven by women, who are more likely to take probiotics than men^2,68^, overall consumer interest is increasing, including about the effectiveness of, and side effects associated with, probiotic consumption^1,2^. This is especially important for women of childbearing age, who may choose to take probiotics during pregnancy without being aware of the safety of these products. Further, clinicians would like to know more about probiotic products^25,26^ to translate this knowledge to their patients and aid them in making evidence-informed clinical decisions. Our evidence review and meta-analysis of the available data suggest a preliminary set of recommendations for research and clinical consideration:

- Researchers should provide detailed and specific safety and adverse effects reporting plans prior to conducting a study, including information and data on:
  - Intervention characteristics: strain type, dosage, duration (e.g. relative to gestational age)
  - Participant information: age, underlying health conditions, diet
  - Adverse effects: type, timing, symptoms, severity, duration, withdrawal due to intervention
- Results of studies, including detailed efficacy and safety data, should be published using accessible language and easy to interpret visuals that can be used by diverse audiences
- Clinicians and consumers should refer to available scientific evidence^61^ on efficacy of probiotic products and their associated adverse effects when making decisions about their use or impact on patient health, and clinicians should disseminate safety information to patients accordingly

Care of prepregnant and pregnant people using or intending to use probiotics, prebiotics or synbiotics can be better managed if we know these products are safe, and the potential adverse effects can be prevented or managed with lifestyle modifications. Prior to this review, comprehensive knowledge on adverse effects associated with probiotic and prebiotic intake in pregnancy and in the postpartum/lactation periods was limited. Scarce and ineffective knowledge translation further limits evidence-informed decision-making about probiotic and prebiotic use, which may impact patient care and an individual’s health. Our study provides a knowledge translation tool for reporting safety outcomes associated with probiotic and prebiotic use in vulnerable populations that are highly receptive to these products. Our analyses suggest that probiotics and prebiotics are safe to use during and after pregnancy and during lactation.

## Data Availability

N/A

## Acknowledgments

This research received no specific grant from any funding agency, commercial or not-for-profit sectors.

## Author Contributions and Notes

H.S. and K.L.C. conceptualised the ideas presented in this paper, planned the methodology, and investigated the evidence. H.S. analysed the data and drafted the manuscript with K.L.C. Both authors contributed to the review and revision of the final paper. The authors declare no conflict of interest.

## Notes

### Competing Interest Statement

The authors have declared no competing interest.

