## Supplementary Files for "Are probiotics and prebiotics safe for use during pregnancy and lactation? A systematic review and meta-analysis"

**Supplementary Table 1.** Preferred Reporting Items for Systematic Review and Meta-Analysis (PRISMA). This checklist was used as a guideline to determine eligibility criteria, information sources and search strategy, data collection and analysis, and risk of bias assessments and data synthesis.

| Section/topic | # | Checklist item | Reported on page # |
|---|---|---|---|
| TITLE | | | |
| Title | 1 | Identify the report as a systematic review, meta-analysis, or both. | 1 |
| ABSTRACT | | | |
| Structured summary | 2 | Provide a structured summary including, as applicable: background; objectives; data sources; study eligibility criteria, participants, and interventions; study appraisal and synthesis methods; results; limitations; conclusions and implications of key findings; systematic review registration number. | 1 |
| INTRODUCTION | | | |
| Rationale | 3 | Describe the rationale for the review in the context of what is already known. | 1 |
| Objectives | 4 | Provide an explicit statement of questions being addressed with reference to participants, interventions, comparisons, outcomes, and study design (PICOS). | 2 |
| METHODS | | | |
| Protocol and registration | 5 | Indicate if a review protocol exists, if and where it can be accessed (e.g., Web address), and, if available, provide registration information including registration number. | N/A |
| Eligibility criteria | 6 | Specify study characteristics (e.g., PICOS, length of follow-up) and report characteristics (e.g., years considered, language, publication status) used as criteria for eligibility, giving rationale. | 2 & Fig S1 |
| Information sources | 7 | Describe all information sources (e.g., databases with dates of coverage, contact with study authors to identify additional studies) in the search and date last searched. | 2 |
| Search | 8 | Present full electronic search strategy for at least one database, including any limits used, such that it could be repeated. | 2 |
| Study selection | 9 | State the process for selecting studies (i.e., screening, eligibility, included in systematic review, and, if applicable, included in the meta-analysis). | 2 |
| Data collection process | 10 | Describe method of data extraction from reports (e.g., piloted forms, independently, in duplicate) and any processes for obtaining and confirming data from investigators. | 2 |
| Data items | 11 | List and define all variables for which data were sought (e.g., PICOS, funding sources) and any assumptions and simplifications made. | 2 |
| Risk of bias in individual studies | 12 | Describe methods used for assessing risk of bias of individual studies (including specification of whether this was done at the study or outcome level), and how this information is to be used in any data synthesis. | 3 |
| Summary measures | 13 | State the principal summary measures (e.g., risk ratio, difference in means). | 3 |
| Synthesis of results | 14 | Describe the methods of handling data and combining results of studies, if done, including measures of consistency (e.g., $I^2$) for each meta-analysis. | 3 & Fig S3 |

| Section/topic | # | Checklist item | Reported on page # |
|---|---|---|---|
| Risk of bias across studies | 15 | Specify any assessment of risk of bias that may affect the cumulative evidence (e.g., publication bias, selective reporting within studies). | 3 & 4 |
| Additional analyses | 16 | Describe methods of additional analyses (e.g., sensitivity or subgroup analyses, meta-regression), if done, indicating which were pre-specified. | N/A |
| **RESULTS** | | | |
| Study selection | 17 | Give numbers of studies screened, assessed for eligibility, and included in the review, with reasons for exclusions at each stage, ideally with a flow diagram. | 3 & Fig S1 |
| Study characteristics | 18 | For each study, present characteristics for which data were extracted (e.g., study size, PICOS, follow-up period) and provide the citations. | 3, 4, 5, Fig 4 & Table S4 |
| Risk of bias within studies | 19 | Present data on risk of bias of each study and, if available, any outcome level assessment (see item 12). | 4 & Fig 1 |
| Results of individual studies | 20 | For all outcomes considered (benefits or harms), present, for each study: (a) simple summary data for each intervention group (b) effect estimates and confidence intervals, ideally with a forest plot. | 4, 5 & Fig 4 |
| Synthesis of results | 21 | Present results of each meta-analysis done, including confidence intervals and measures of consistency. | 5 & Fig 4 |
| Risk of bias across studies | 22 | Present results of any assessment of risk of bias across studies (see Item 15). | 4 & Fig 1 |
| Additional analysis | 23 | Give results of additional analyses, if done (e.g., sensitivity or subgroup analyses, meta-regression [see Item 16]). | 5 & Fig S3 |
| **DISCUSSION** | | | |
| Summary of evidence | 24 | Summarize the main findings including the strength of evidence for each main outcome; consider their relevance to key groups (e.g., healthcare providers, users, and policy makers). | 7, 8 |
| Limitations | 25 | Discuss limitations at study and outcome level (e.g., risk of bias), and at review-level (e.g., incomplete retrieval of identified research, reporting bias). | 8 |
| Conclusions | 26 | Provide a general interpretation of the results in the context of other evidence, and implications for future research. | 8 |
| **FUNDING** | | | |
| Funding | 27 | Describe sources of funding for the systematic review and other support (e.g., supply of data); role of funders for the systematic review. | N/A |

**Supplementary Table 2.** Classification for primary outcome groups.

| Primary Outcome Group | Definition |
|---|---|
| Maternal microbiome and gastrointestinal health | Involving the maternal microbiota at multiple body sites, including vaginal and gut microbiomes, and gastrointestinal symptoms, such as constipation. |
| Maternal metabolic health | Involving maternal insulin metabolism, plasma glucose, glycemic status, gestational diabetes mellitus, overweight and obesity, anthropometric measurements, and serum lipid and leptin concentrations. |
| Maternal mental health | Involving maternal mental health, depression and anxiety. |
| Maternal breastmilk and breast health | Involving infectious mastitis, breastmilk composition, breastmilk microbiota and immunomodulatory effects in lactation. |
| Infant microbiome and gastrointestinal health | Involving the infant gut microbiome and gastrointestinal symptoms, such as diarrhea. |
| Infant metabolic health | Involving infant blood pressure and metabolic programming and child blood lipid concentrations. |
| Infant allergy and immune health | Involving infant eczema and other allergic disease. |
| Infant growth and behaviour | Involving infant health outcomes (birthweight, disease symptoms and onset), growth and sleep duration. |
| Pregnancy outcome | Involving pregnancy rate post-*in vitro* fertilization, incidence and prevention of preterm delivery, incidence of pre-eclampsia and premature membrane rupturing. |

**Supplementary Table 3.** Absolute risk difference (ARD) for studies (*n*=11) with reported adverse effects.

| Study | Adverse Effect Type | Absolute Risk Difference (95% CI) |
|---|---|---|
| Asgharian, 2020 | • Stomachache | 0 (-4.30, 4.30) |
| Gille, 2016 | • Increased vaginal discharge<br>• Changes in stool consistence | -5.00 (-9.44, -0.55) |
| Mirghafourvand, 2016 | • Nausea<br>• Bloating<br>• Diarrhea | 3.30 (-21.8, 28.5) |
| Boyle, 2011 | • Gastrointestinal symptoms, unspecified | 8.00 (-1.86, 17.9) |
| Krauss-Silva, 2011 | • Nausea<br>• Vomiting<br>• Diarrhea<br>• Sleepiness<br>• Headache<br>• Pyrosis<br>• Stomachache<br>• Nausea/vomiting and headache<br>• Vomiting and diarrhea<br>• Pruritus<br>• Abdominal pain/fainting<br>• Abdominal ache<br>• Abdominal pain/vomiting<br>• Bloody diarrhea<br>• Tachycardia | 1.00 (-3.47, 5.57) |
| Shadid, 2007 | • Constipation and bloating<br>• Diarrhea or reflux | 6.60 (-13.0, 26.3) |
| Pellonpera, 2019 | • Gastrointestinal symptoms, unspecified<br>• Headache | -2.10 (-13.9, 9.80) |
| Mantaring, 2018 | • Maternal nausea, vomiting and diarrhea<br>• Infantile serious diarrhea<br>• Infantile diarrhea | -13.8 (-27.9, 0.37) |
| Rautava, 2012 | • Maternal gastrointestinal symptoms, unspecified<br>• Maternal aggravation of eczema<br>• Infantile gastrointestinal symptoms, unspecified | -12.5 (-32.0, 7.08) |
| | • Maternal gastrointestinal symptoms, unspecified<br>• Infantile gastrointestinal symptoms, unspecified | -1.90 (-21.6, 17.9) |
| Arroyo, 2010 | • Flatulence | -7.3 (-11.8, -2.69) |
| Laitinen, 2009 | • Flatulence, loose stools or constipation | -4.00 (-10.9, 3.07) |

A negative ARD signifies increased risk of the adverse effect(s), whereas a positive ARD signifies decreased risk of the adverse effect(s).

**Supplementary Table 4.** Full citations of studies reporting adverse effects (*n*=11).

| Study name | Full citation |
|---|---|
| Asgharian, 2020 | Asgharian, H., Homayouni-Rad, A., Mirghafourvand, M., Mohammad-Alizadeh-Charandabi, S. Effect of probiotic yoghurt on plasma glucose in overweight and obese pregnant women: a randomized controlled clinical trial. *European Journal of Nutrition* **2020**, *59*, 205-215, doi:10.1007/s00394-019-01900-1. |
| Gille, 2016 | Gille, C., Boer, B., Marschal, M., Urschitz, M. S., Heinecke, V., Hund, V., Speidel, S., Tarnow, I., Mylonas, I., Franz, A., Engel, C., Poets, C. F. Effect of probiotics on vaginal health in pregnancy. EFFPRO, a randomized controlled trial. *American Journal of Obstetrics and Gynecology* **2016**, *215*, doi:10.1016/j.ajog.2016.06.021. |
| Mirghafourvand, 2016 | Mirghafourvand, M.R., A. H. Alizadeh, S. M. Fardiazar, Z. Shokri, K. The Effect of Probiotic Yogurt on Constipation in Pregnant Women: A Randomized Controlled Clinical Trial. *Iranian Red Crescent Medical Journal* **2016**, *18*, doi:10.5812/ircmj.39870. |
| Boyle, 2011 | Boyle, R.J., Ismail, I. H., Kivivuori, S., Licciardi, P. V., Robins-Browne, R. M., Mah, L. J., Axelrad, C.. Moore, S., Donath, S., Carlin, J. B., Lahtinen, S. J., Tang, M. L. K. Lactobacillus GG treatment during pregnancy for the prevention of eczema: a randomized controlled trial. *Allergy* **2011**, *66*, 509-516, doi:10.1111/j.1398-9995.2010.02507.x. |
| Krauss-Silva, 2011 | Krauss-Silva, L.M., M. E. L. Alves, M. B. Braga, A. Camacho, K. G. Batista, M. R. R. Almada-Horta, A. Rebello, M. R. Guerra, F. A randomised controlled trial of probiotics for the prevention of spontaneous preterm delivery associated with bacterial vaginosis: preliminary results. *Trials* **2011**, *12*, doi:10.1186/1745-6215-12-239. |
| Shadid, 2007 | Shadid, R.H., M. Knol, J. Theis, W. Beermann, C. Rjosk-Dendorfer, D. Schendel, D. J. Koletzko, B. V. Krauss-Etschmann, S. Effects of galactooligosaccharide and long-chain fructooligosaccharide supplementation during pregnancy on maternal and neonatal microbiota and immunity - a randomized, double-blind, placebo-controlled study. *American Journal of Clinical Nutrition* **2007**, *86*, 1426-1437. |
| Pellonpera, 2019 | Pellonpera, O.M., K. Houttu, N. Vahlberg, T. Koivuniemi, E. Tertti, K. Ronnemaa, T. Laitinen, K. Efficacy of Fish Oil and/or Probiotic Intervention on the Incidence of Gestational Diabetes Mellitus in an At-Risk Group of Overweight and Obese Women: A Randomized, Placebo-Controlled, Double-Blind Clinical Trial. *Diabetes Care* **2019**, *42*, 1009-1017, doi:10.2337/dc18-2591. |
| Mantaring, 2018 | Mantaring, J.B., J. Destura, R. Pecquet, S. Vidal, K. Volger, S. Guinto, V. Effect of maternal supplement beverage with and without probiotics during pregnancy and lactation on maternal and infant health: a randomized controlled trial in the Philippines. *Bmc Pregnancy and Childbirth* **2018**, *18*, doi:10.1186/s12884-018-1828-8. |
| Rautava, 2012 | Rautava, S.K., E. Salminen, S. Isolauri, E. Maternal probiotic supplementation during pregnancy and breast-feeding reduces the risk of eczema in the infant. *Journal of Allergy and Clinical Immunology* **2012**, *130*, 1355-1360, doi:10.1016/j.jaci.2012.09.003. |
| Arroyo, 2010 | Arroyo, R., Martín, V., Maldonado, A., Jiménez, E., Fernández, L., Rodríguez, J. M. Treatment of infectious mastitis during lactation: antibiotics versus oral administration of Lactobacilli isolated from breast milk. *Clin Infect Dis* **2010**, *50*, 1551-1558, doi:10.1086/652763. |
| Laitinen, 2009 | Laitinen, K.P., T. Isolauri, E. Nutr Allergy Mucosal Immunology, In. Probiotics and dietary counselling contribute to glucose regulation during and after pregnancy: a randomised controlled trial. *British Journal of Nutrition* **2009**, *101*, 1679-1687, doi:10.1017/s0007114508111461. |

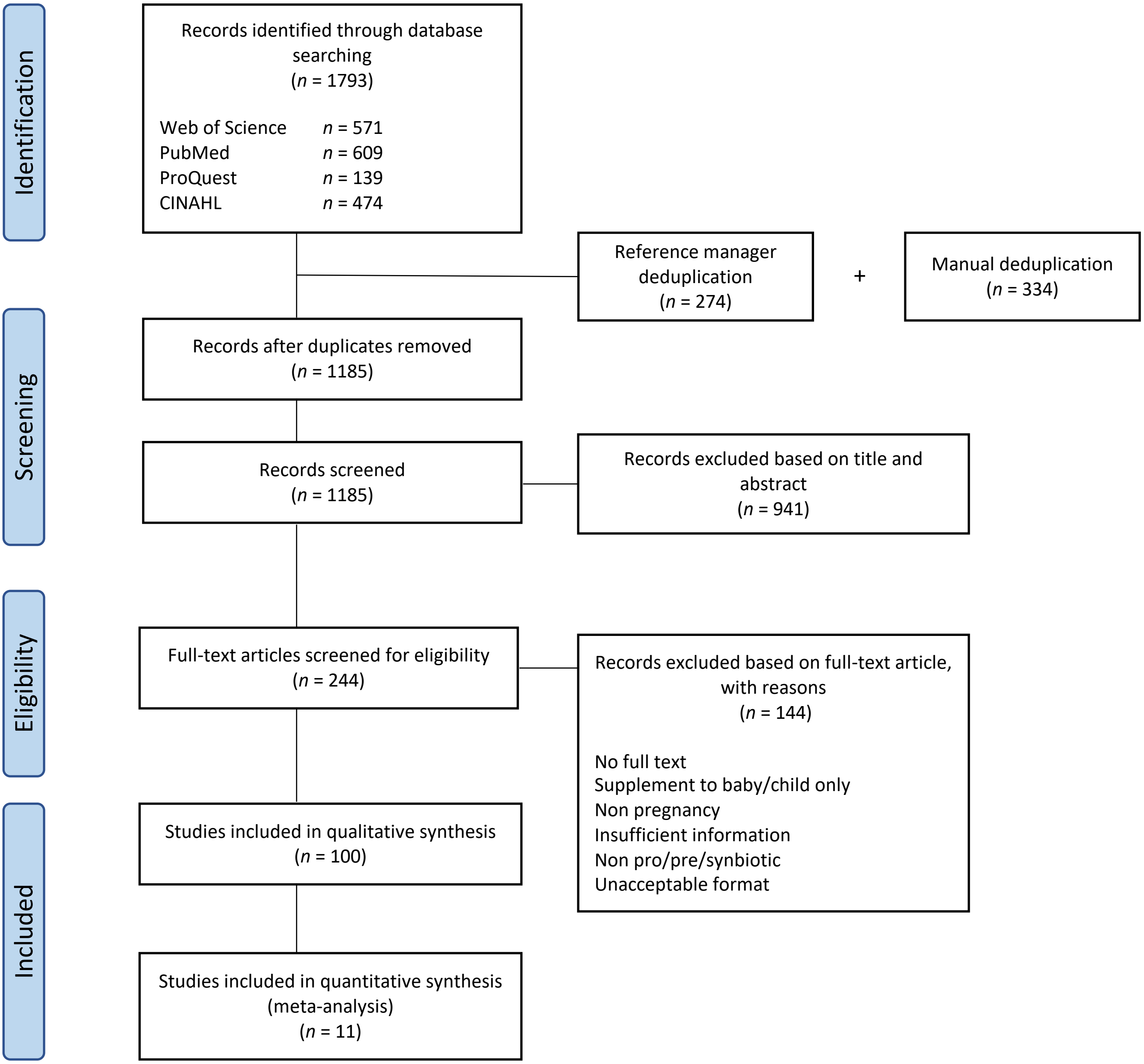


**Supplementary Figure 1.** PRISMA flow diagram for article selection and review.

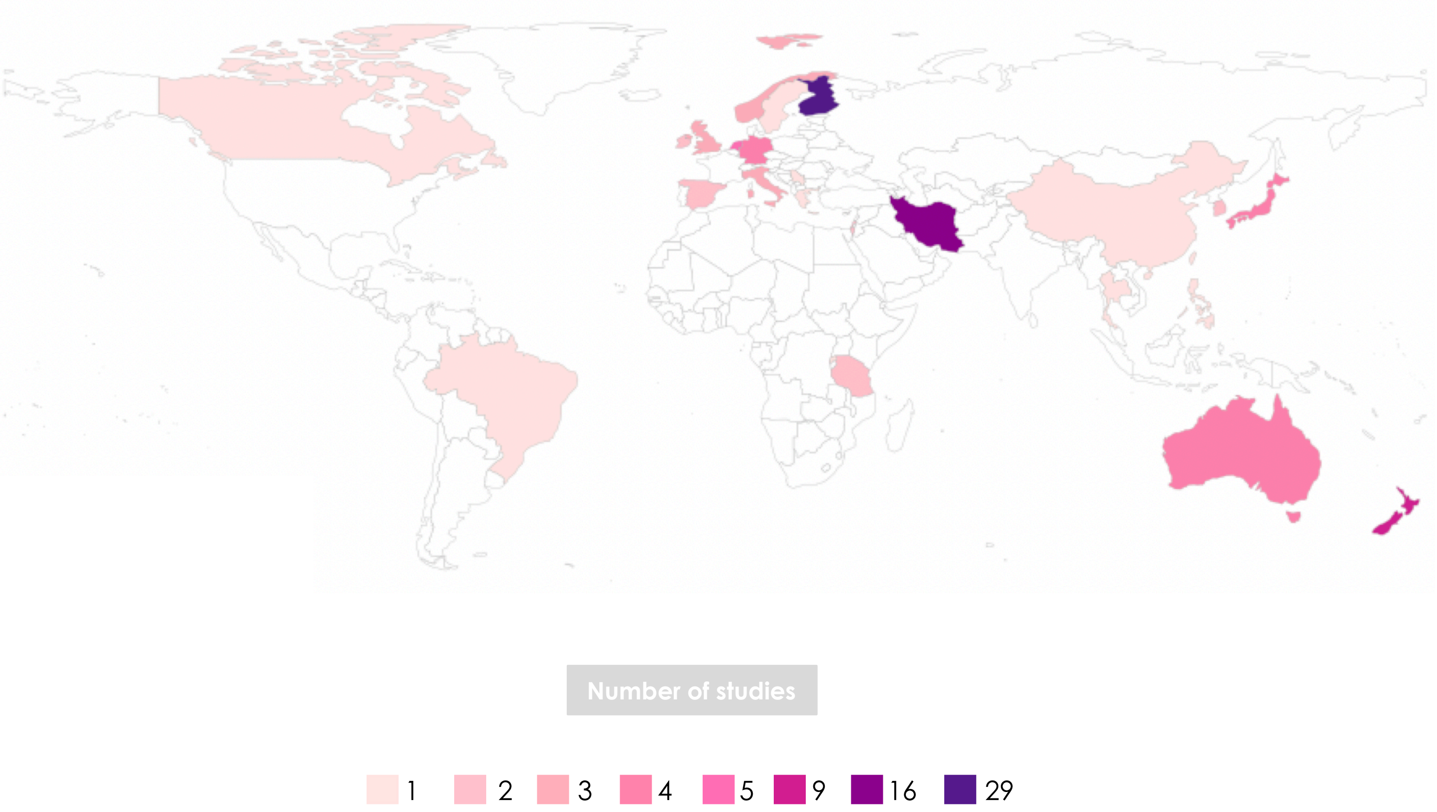


**Supplementary Figure 2.** World heatmap illustrating geographical location of each study included in the review. Data are shown for 100 studies from 25 countries. Increasing study counts are represented by darker colours. Fifty-four percent of studies were from Europe (*n*=54), 28% from Asia (*n*=28), 13% from Oceania (*n*=13), and the remaining 6% from North America, South America and Africa (*n*=6). Finland had the highest representation in studies under review, representing 29% of all studies (*n*=29).

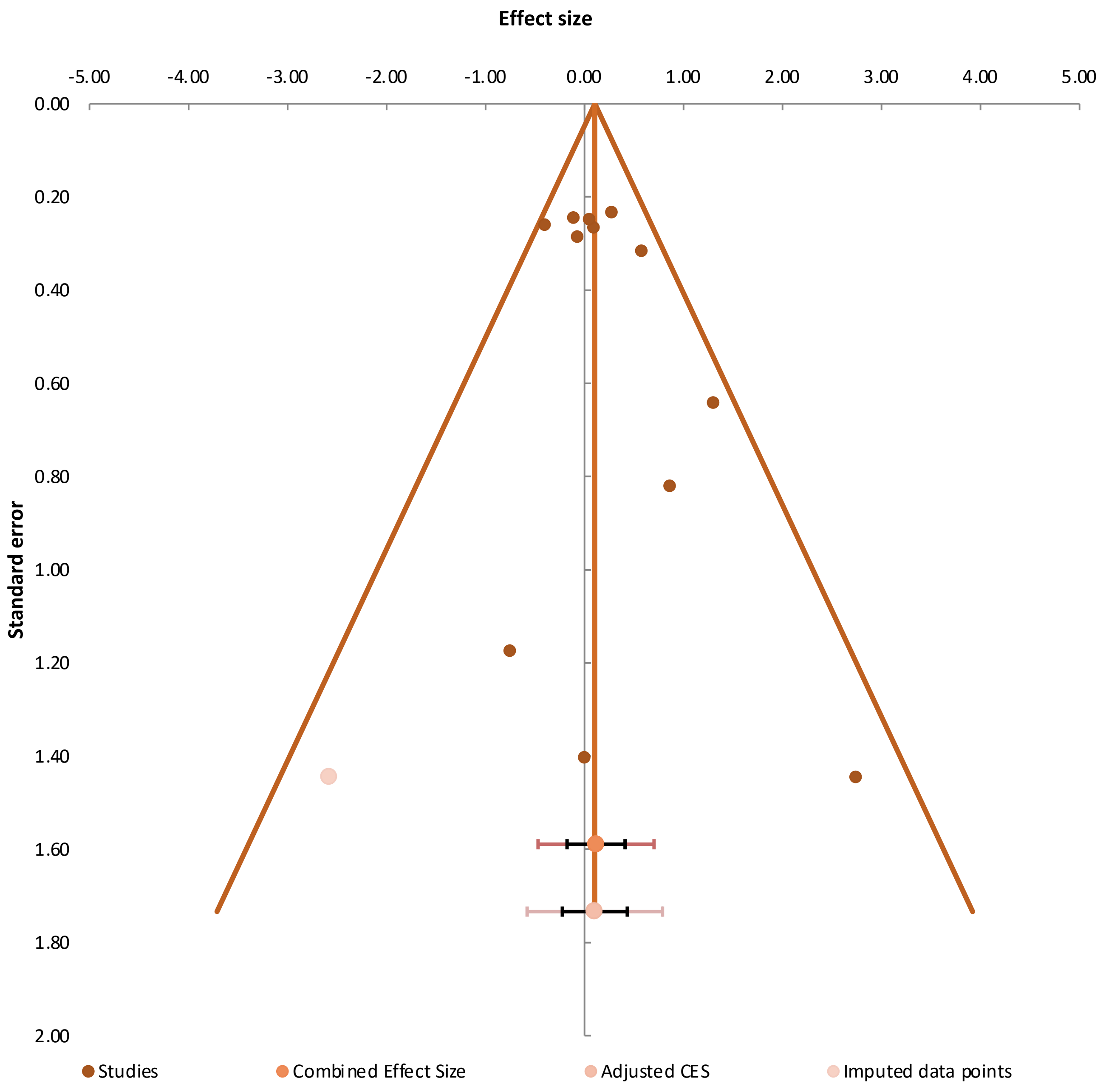


**Supplementary Figure 3.** Funnel plot of log risk ratio values against standard error for each study with reported adverse effects (*n* = 11 studies). One study (Rautava et al.), with two intervention groups and one control group, was treated as two individual studies (thus two dots are represented on the funnel plot) to account for adverse effects that were reported in both intervention groups. The vertical line represents the summary effect size estimates (log risk ratio), and the slanted lines are pseudo 95% CIs. The combined effect size (CSE) is the log risk ratio of all studies combined. The adjusted combined effect size and accompanying confidence and prediction intervals in this plot represent the results of a trim-and-fill procedure. These adjusted statistics are based upon the set of initially included studies expanded with the imputed data points. Funnel plot shows little asymmetry.
